# The D614G virus mutation enhances anosmia in COVID-19 patients: Evidence from a systematic review and meta-analysis of studies from South Asia

**DOI:** 10.1101/2021.08.11.21261934

**Authors:** Christopher S. von Bartheld, Molly M. Hagen, Rafal Butowt

**Affiliations:** Department of Physiology and Cell Biology, University of Nevada, Reno School of Medicine, Reno, Nevada 89557, USA; School of Public Health, University of Nevada, Reno, Nevada, USA; L. Rydygier Collegium Medicum, Nicolaus Copernicus University, 85-094 Bydgoszcz, Poland

**Keywords:** COVID-19, anosmia prevalence, olfactory dysfunction, SARS-CoV-2, D614G virus mutation, South Asia

## Abstract

The prevalence of chemosensory dysfunction in patients with COVID-19 varies greatly between populations. It is unclear whether such differences are due to factors at the level of the human host, or at the level of the coronavirus, or both. At the host level, the entry proteins which allow virus binding and entry have variants with distinct properties, and the frequency of such variants differs between ethnicities. At the level of the virus, the D614G mutation enhances virus entry to the host cell. Since the two virus strains (D614 and G614) co-existed in the first six months of the pandemic in most populations, it has been difficult to distinguish between contributions of the virus and contributions of the host for anosmia. To answer this question, we conducted a systematic review and meta-analysis of studies in South Asian populations when either the D614 or the G614 virus was dominant. We show that populations infected predominantly with the G614 virus had a much higher prevalence of anosmia (pooled prevalence of 31.8%) compared with the same ethnic populations infected mostly with the D614 virus strain (pooled anosmia prevalence of 5.3%). We conclude that the D614G mutation is a major contributing factor that increases the prevalence of anosmia in COVID-19, and that this enhanced effect on olfaction constitutes a previously unrecognized phenotype of the D614G mutation. The new virus strains that have additional mutations on the background of the D614G mutation can be expected to cause a similarly increased prevalence of chemosensory dysfunctions.

**Graphical Abstract:** 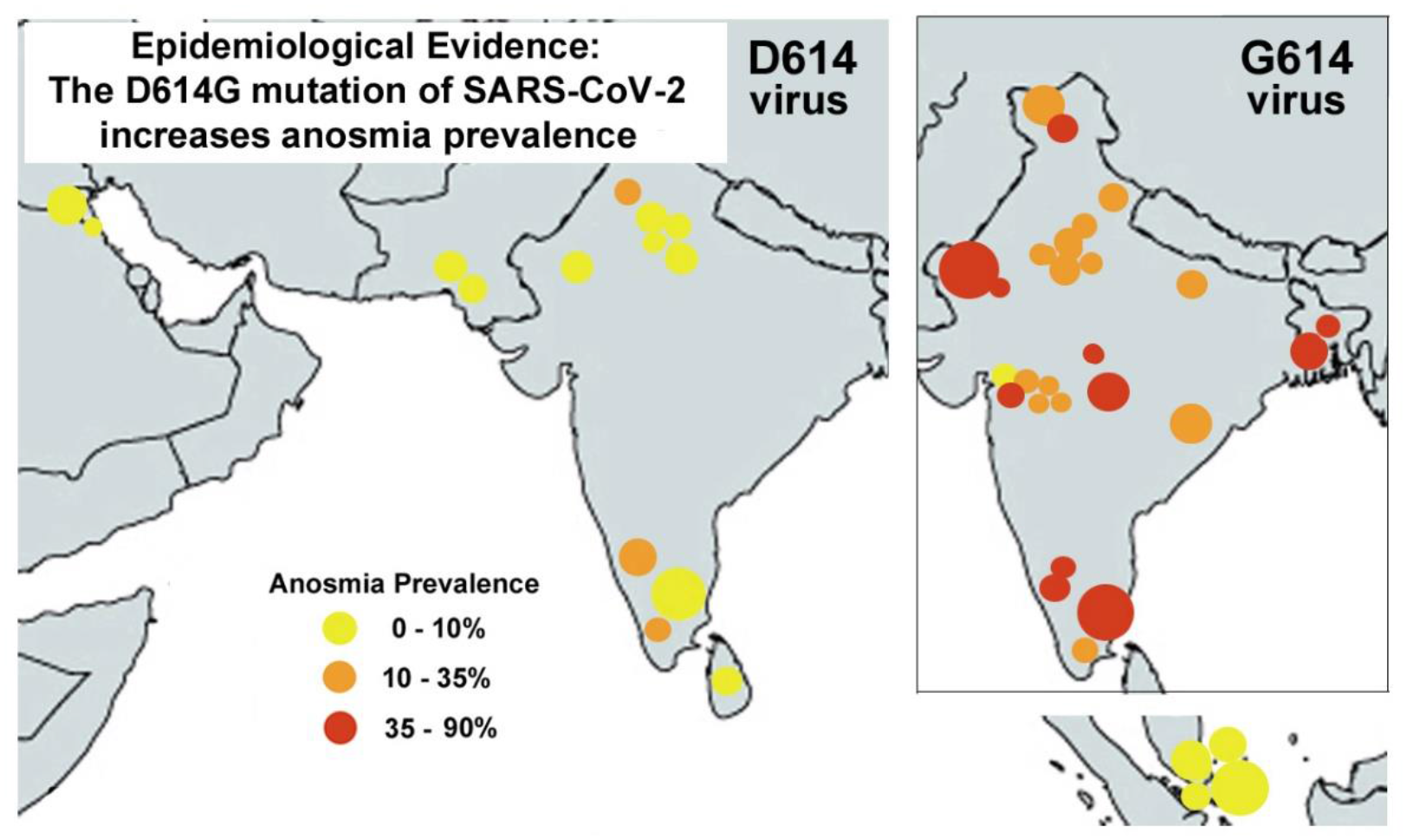

## Introduction

Chemosensory dysfunction has been identified as one of the most frequent symptoms of COVID-19.^1–4^ However, the prevalence of smell loss in COVID-19 varies greatly between populations. Some studies reported the prevalence of anosmia or hyposmia to be lower than 1%, while others reported a prevalence of over 70%.^1, 2, 4–7^ Such differences may be due to multiple factors, but two reasons are currently considered to be most relevant: differences at the level of the human host (variants in the virus entry proteins, angiotensin-converting enzyme 2, ACE2, and/or in the protease TMPRSS2), and differences at the level of the virus (mutations of the spike protein resulting in an altered efficiency of entry to the host cell, and therefore higher infectivity), or a combination of these host and virus factors. ^1, 2, 4, 5, 8–13^

For historical reasons of how the pandemic unfolded, it has been difficult to define the relative contributions of host and virus factors for anosmia. The pandemic started in East Asia with the less infectious D614 virus strain, but when COVID-19 reached Europe and North America and the rest of the world, the virus with a D614G mutation in the spike protein rapidly replaced the original D614 virus. ^14^ Since the pandemic was largely controlled in East Asia in the second half of 2020, there are very few studies from East Asia on prevalence of anosmia after the initial D614 virus infestation. Studies reporting anosmia prevalence from the rest of the world typically collected data from regions where the two virus strains co-existed or when the G614 virus was dominant. ^1, 4, 11, 14^ This made it impossible to discern whether the virus mutation or host protein variants are primarily responsible for the differences in anosmia prevalence between populations in East Asia and the rest of the world.

Here, we took advantage of the fact that early studies from South Asia collected data on anosmia prevalence in regions with 50% or more D614 virus infections, while more recent studies collected data in South Asia when the G614 strain was dominant. Our systematic review and meta-analysis of the same ethnicity (South Asians) shows a strong association between the D614 virus and low prevalence of anosmia, and a strong association between the G614 virus and high anosmia prevalence. The most parsimonious interpretation of these data is that the D614G mutation plays a significant, and apparently the most important, role in causing an increased prevalence of anosmia in COVID-19 patients. We here present our evidence and we discuss the presumed underlying molecular mechanisms of how SARS-CoV-2, and especially the G614 virus, causes olfactory dysfunction.

### The spread and increasing dominance of the G614 virus over the D614 virus

In the beginning of the pandemic, the D614 virus emerged in China and spread to the rest of Asia, Europe, the Middle East, Australia, and North and South America. The spread beyond East Asia was accompanied by the appearance of the more infectious G614 virus mutation, and over the next weeks and months this strain became dominant over the original D614 strain in most parts of the world. ^11, 14, 15^ The D614 virus remained dominant beyond May of 2020 in only few regions: most of China, Singapore, Malaysia, the South of India and the Delhi region (https://www.gisaid.org/; https://cov.lanl.gov/apps/covid-19/map/) ^14, 16–25^ The co-existence of the D614 and G614 viruses made it impossible, in most regions, to discern whether differences in the prevalence of anosmia were due to differences among the host populations (frequency of variants of the entry proteins ACE2 or TMPRSS2 and thus ethnic differences), or were due to differences in the infectivity and cell entry efficiencies of the coronavirus. In East Asia, nearly all studies were conducted when the D614 virus was dominant. In Europe, the Americas, the Middle East and Africa, there was either a co-existence of D614 and G614 viruses, or a dominance of the G614 virus during the periods of data collection for anosmia prevalence. To clarify the suspected role of the virus type for anosmia, it was necessary to find an ethnicity which was infected either at different times or in different regions when either the D614 or G614 virus was dominant, and to compare the prevalence of anosmia between studies reporting on such cohorts. There are no suitable and unambiguous (unequivocal) pairings of appropriate studies from Europe, the Middle East, the Americas or East Asia that allow comparison of the same ethnicity for the two types of virus strains. Fortunately, we found that one ethnicity does meet these criteria – South Asians.

### Spatiotemporal mapping of D614 or G614 virus dominance in South Asians

The spread of the SARS-CoV-2 virus into the Indian subcontinent began at the end of January 2020, with waves from several regions: One from South East Asia (and the Middle East), carrying the D614 virus, and another wave primarily from Europe and the USA, carrying the G614 virus. Overall in India, the D614 virus dominated until late April or May, while the G614 virus became dominant in most Indian states mid to late May or June 2020. ^16, 17, 19–21, 27, 28^ However, there were significant regional differences. For example, the D614 virus persisted for a longer time in the Delhi region, ^17, 19^ and also in the South of India, ^17, 20, 21, 24, 27–30^ while the G614 virus dominated early in the West of India (Gujarat, Maharashtra), ^17, 19, 25, 31^ and also in the Eastern and Central Indian states ^28^ as well as in Bangladesh. ^14, 26, 32, 33^ Pakistan and Sri Lanka had different waves of virus spread, with an early D614 dominance, but becoming all G614 in June and July 2020. ^34–36^ Importantly, the predominance of the G614 virus was delayed in South India until mid May to June of 2020. ^17, 18, 20–22, 25, 30^ Such regional differences were caused by travel routes as well as regional spreader events, e.g., in Delhi. ^17^ Indians and Bangladeshis were exposed primarily to the D614 virus in March through June or July of 2020 in some regions outside the Indian subcontinent, e.g., Kuwait, ^37^ Malaysia, ^38, 39^ and Singapore, ^14, 23^ while South Asians in Oman were mostly exposed to the G614 virus in March and April of 2020. ^37, 40^ The temporospatial distribution of the D614 virus vs G614 virus in South Asia is summarized in Fig. 1A-F.

**Fig. 1A-F.**
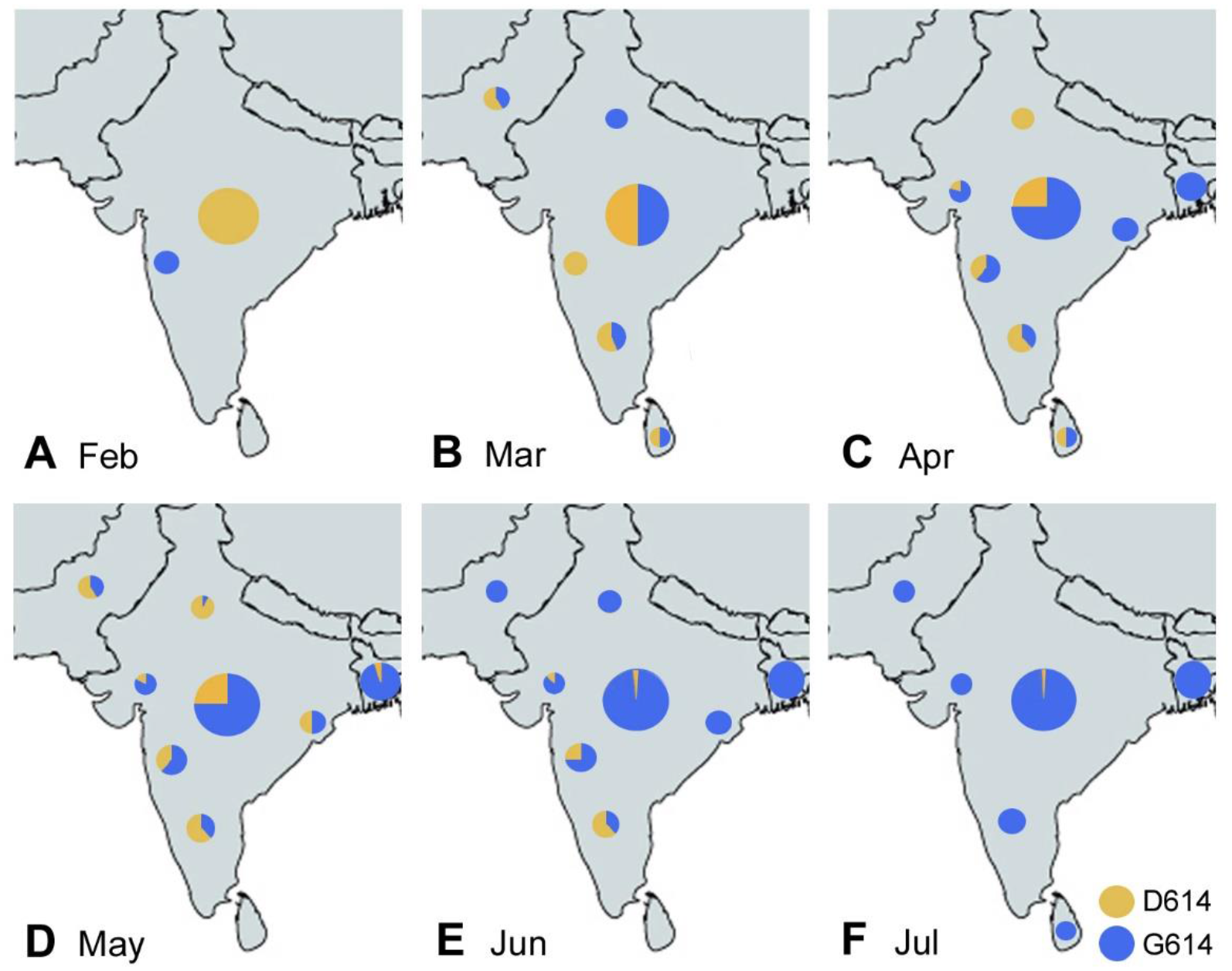
Map of South Asia showing contributions of D614 and G614 virus to COVID-19 in February to July of the year 2020. Virus strain dominance according to relevant references. ^14, 16–22, 24–29, 31–36^ The large circle in the center of India shows the overall contributions of D614 and G614, regardless of region; the small circles represent regional contributions of D614 and G614 during the indicated month of 2020.

### Systematic review and meta-analysis of South Asian studies

We here explore the hypothesis that there was a lower anosmia prevalence when the D614 virus was the prevailing strain, while a higher anosmia prevalence was induced by the G614 virus. Therefore, we searched the literature and conducted a systematic review and meta-analysis on studies reporting olfactory dysfunction in South Asian patients with COVID-19, sorted by dominance of the two virus strains. Dominance of D614 vs G614 virus was determined by reviewing studies that mapped the temporospatial changes within different regions of the Indian subcontinent. ^17, 19, 25, 28, 33–35^ For our systematic review and search strategy, we adhered to the PRISMA guidelines ^41^ (Fig. 2).

**Fig. 2:**
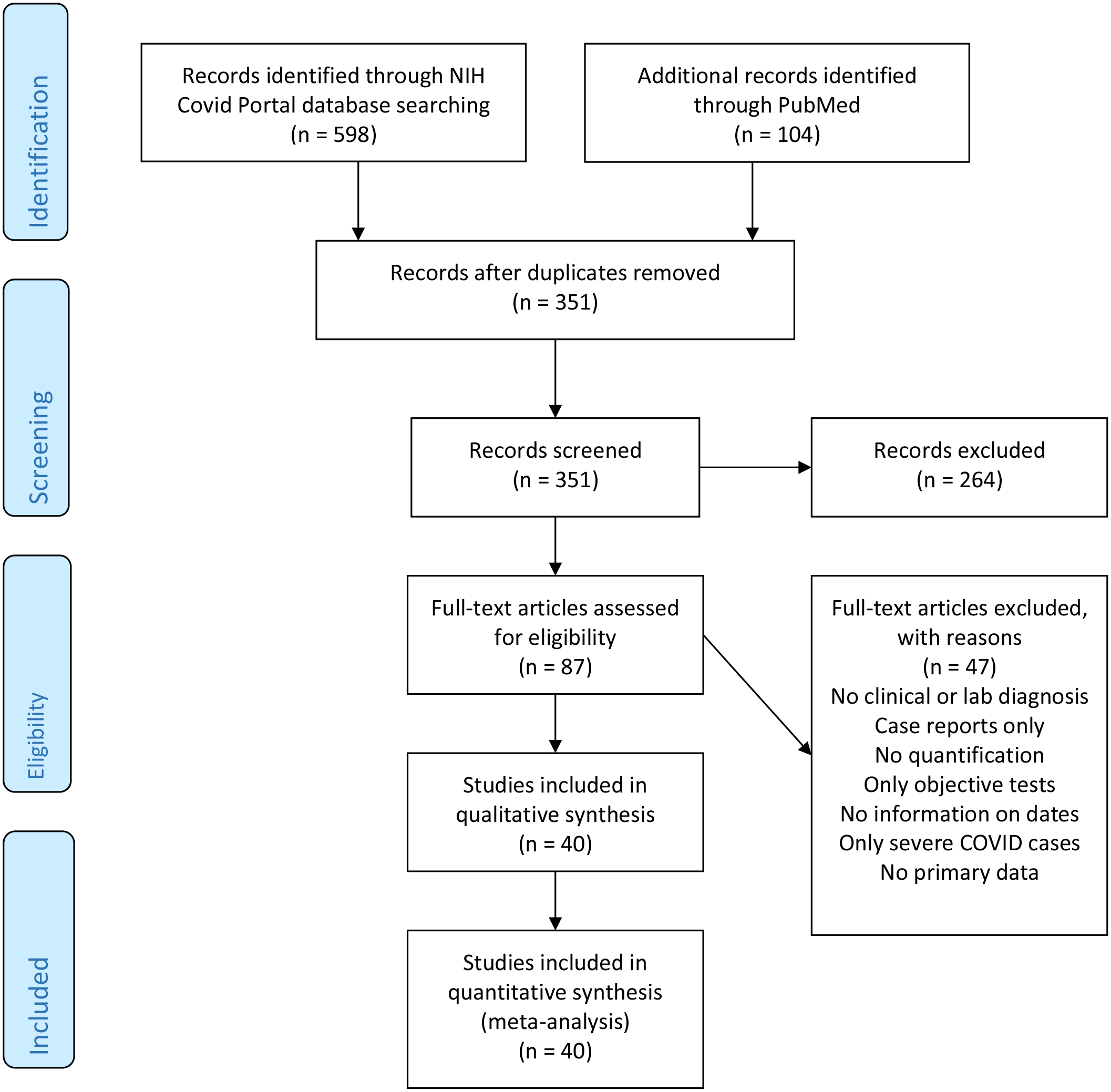
Flow chart of literature search and systematic review of studies reporting on COVID-19 related olfactory dysfunction in South Asians through July 22, 2021.

Our search retrieved 598 studies that examined South Asians for COVID-related loss of smell, of which 40 met the inclusion criteria (see Methods). These 40 studies (reporting on 43 cohorts) were subjected to a meta-analysis (Table 1). Fifteen studies reported anosmia prevalence in eighteen cohorts with a total of 7,247 COVID-19 patients from regions where the D614 virus was dominant: three studies from Kuwait and Singapore ^42–44^ and twelve from India or Pakistan. ^45–56^ We compared such data with the results obtained in 25 studies reporting on 25 cohorts with a total of 9,626 South Asian patients from the Indian subcontinent (India and Bangladesh) and Oman, when the G614 virus had become dominant (Table 1, illustrated in Fig. 3A, B). ^6, 57–80^ The differences in results between the two types of cohorts are shown in the forest plots (Fig. 4A). The pooled prevalence of olfactory dysfunction in the same ethnicity (South Asians) in regions with D614 predominance was 5.33% (95% confidence interval, CI, = 3.52-8.00%), while in regions with G614 predominance, it was 31.79% (95% CI = 23.26-41.76%) (Fig. 4A, C). The subgroup test from the random effects meta-analysis showed that this was a statistically significant difference with p < 0.0001 (Fig. 4C).

**Fig. 3A, B.**
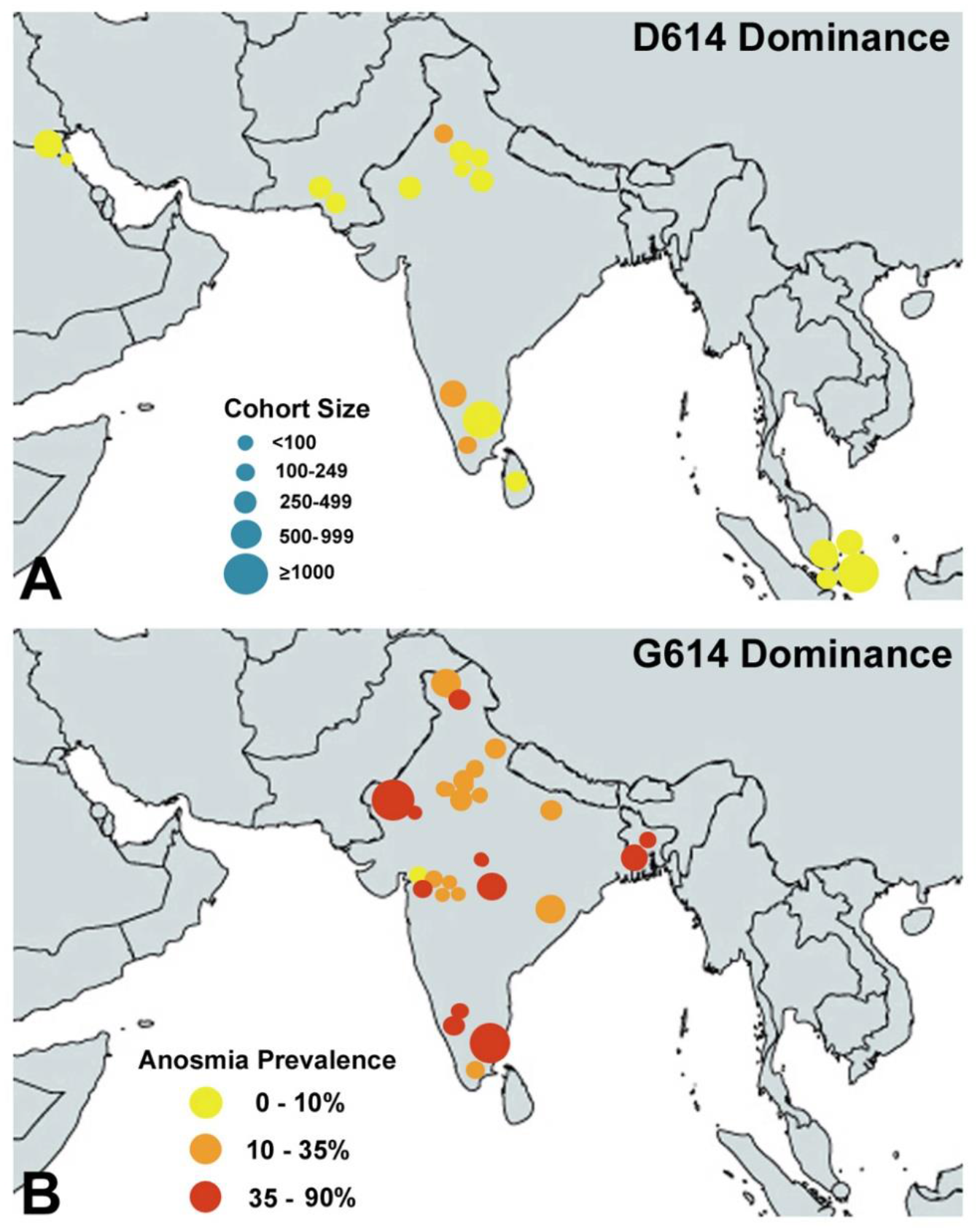
Location of studies reporting the prevalence of olfactory dysfunction among South Asians with D614 virus predominance (A), and G614 virus predominance (B). The cohort size is indicated by the size of the blue dots, the prevalence of olfactory dysfunction is indicated by the heat map, increasing from yellow to red. Note that mostly D614 infections lead rarely to a more than 10% anosmia prevalence, while almost all of the mostly G614 infections lead to a prevalence of 10-90%, in the same ethnicity (South Asians).

**Fig. 4A-D.**
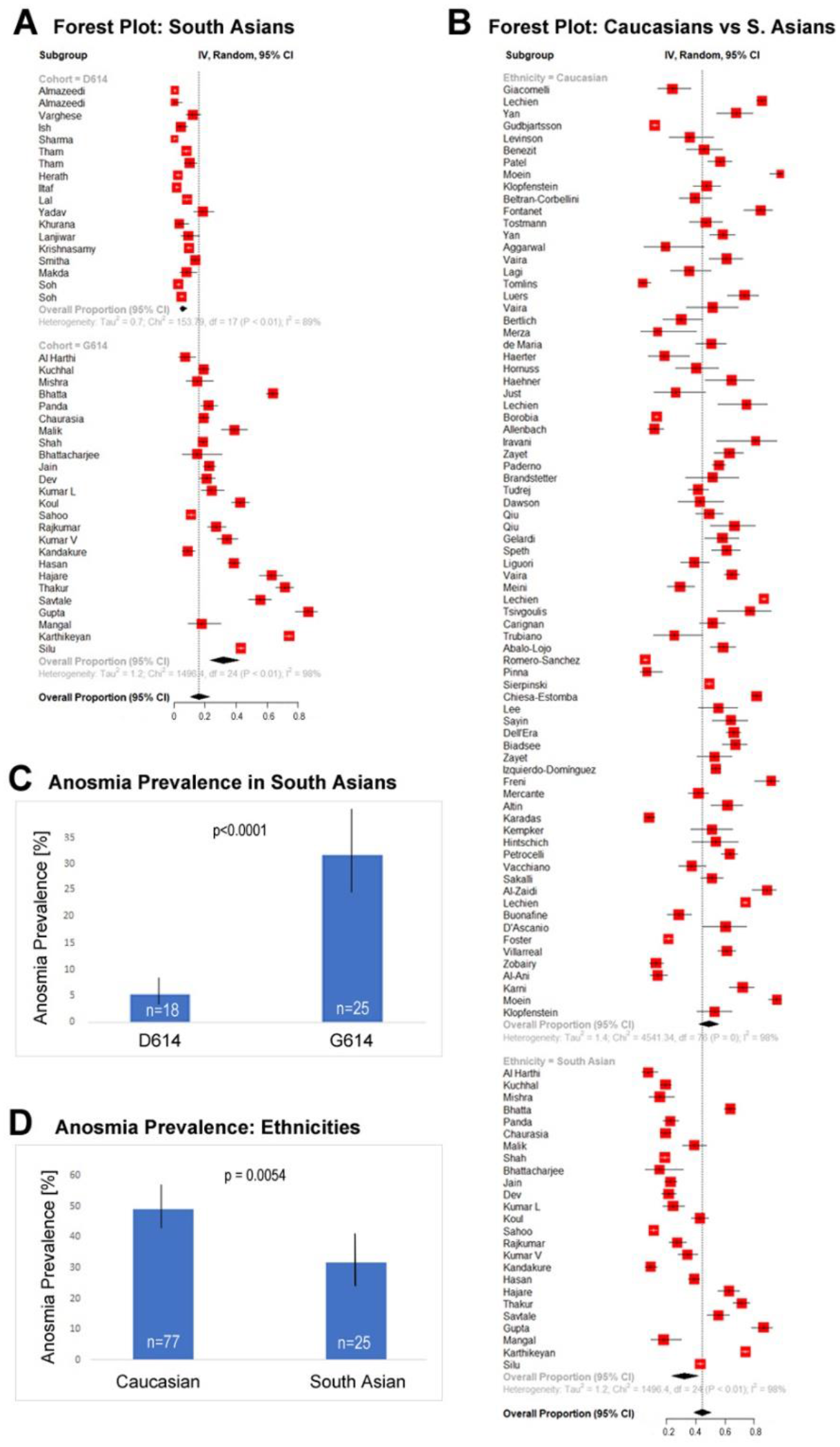
Comparison of the prevalence of olfactory dysfunction in populations infected with D614 or G614 virus predominance. **A.** Forest plots of olfactory dysfunction prevalence in South Asians infected mostly with D614 vs G614 virus. **B.** Forest plots of olfactory dysfunction prevalence in Caucasians vs. South Asians infected mostly with the G614 virus. **C.** Bar graph comparing the pooled anosmia prevalence with 95% confidence intervals and p-value between D614 and G614 cohorts. **D.** Bar graph showing the pooled prevalence of anosmia in Caucasians vs. mostly G614 virus-infected South Asians (Caucasian cohorts from von Bartheld et al., 2020). ^1^ Numbers of cohorts are indicated in white on the bars.

**TABLE 1.** Smell Dysfunction in COVID-19: Chronology of Studies on South Asians with either D614 virus dominance or G614 virus dominance.

| Author | Reference # | Start Data Collection<br>mm dd yy or month and year | End Data Collection<br>mm dd yy or month and year | Ethnicity (Location)<br>North, West, South, East<br>India (N, W, S, E) | Cohort # | Smell Loss Prevalence (%) | Age mean, m median, M | Male Gender (%) | Severe COVID disease (%) |
| --- | --- | --- | --- | --- | --- | --- | --- | --- | --- |
| **D614 VIRUS DOMINANCE** |  |  |  |  |  |  |  |  |  |
| Almazeedi | 42 | 02 24 20 | 04 20 20 | Indian (Kuwait) | 527 | 0.1 | M 41 | 81 | 1.1 |
| Almazeedi | 42 | 02 24 20 | 04 20 20 | Bangladeshi (Kuwait) | 70 | 0.1 | M 41 | 81 | 5.7 |
| Varghese | 45 | Feb 2020 | 03 31 20 | S India (Kerala) | 202 | 12 | m 36.5 | 80.2 | 3 |
| Ish | 46 | 03 01 20 | 04 30 20 | N India (Delhi) | 170 | 4.1 | N D | N D | 0 |
| Sharma | 47 | 03 20 20 | 04 30 20 | N India (Rajasthan) | 234 | 0.0 | m 35.1 | 64.5 | 0 |
| Tham | 43 | 03 24 20 | 04 16 20 | Bangladeshi (Singapore) | 568 | 7.74 | m 34 | 87.6 | N D |
| Tham | 43 | 03 24 20 | 04 16 20 | Indian (Singapore) | 221 | 9.95 | m 34 | 87.6 | N D |
| Herath | 48 | 03 10 20 | 05 30 20 | Sri Lanka | 431 | 2.3 | m 37 | 78 | 1.2 |
| Iltaf | 49 | Mar 2020 | Jun 2020 | Pakistan (Karachi) | 350 | 1.4 | m 49.5 | 70 | N D |
| Lal | 50 | 03 01 20 | 06 30 20 | N India (Delhi) | 435 | 8.0 | m 38.3 | 66.7 | N D |
| Yadav | 51 | 03 01 20 | 07 05 20 | N India (Punjab) | 152 | 18.4 | m 43 | 51.3 | 0 |
| Khurana | 52 | Apr 2020 | Apr 2020 | N India (Delhi) | 94 | 3.2 | m 36 | 59.6 | 1 |
| Lanjiwar | 53 | 04 01 20 | 04 20 20 | N India (Delhi) | 109 | 9.2 | m 38.2 | 86.2 | 0 |
| Krishnasamy | 54 | Apr 2020 | May 2020 | S India (Tamil Nadu) | 1,263 | 9.4 | M 35 | 66.3 | 10.5 |
| Smitha | 55 | Apr 2020 | Jun 2020 | S India (Karnataka) | 500 | 13.4 | ~ 40 | 62 | 5 |
| Makda | 56 | Apr 2020 | Jul 2020 | Pakistan (Karachi) | 114 | 7.8 | m 51 | 54.4 | 54.4 |
| Soh | 44 | 05 01 20 | 07 01 20 | Indian (Singapore) | 743 | 2.6 | m 35 | 100 | 0 |
| Soh | 44 | 05 01 20 | 07 01 20 | Bangladeshi (Singapore) | 1,064 | 4.7 | m 35 | 100 | 0 |
| TOTAL |  |  |  |  | 6,247 | Mean= 5.3% | Mean= 38.83 | Mean= 75.08 | Mean= 5.85% |
| **G614 VIRUS DOMINANCE** |  |  |  |  |  |  |  |  |  |
| Al Harthi | 57 | 03 03 20 | 05 09 20 | South Asian (Oman) | 102 | 6.9 | m 26 | 77.5 | 18.6 |
| Kuchhal | 58 | 04 01 20 | 07 31 20 | N India (Uttarakhand) | 465 | 18.9 | N D | N D | N D |
| Mishra | 59 | Apr 2020 | May 2020 | W India (Maharashtra) | 74 | 14.8 | ~ 35 | 58.1 | N D |
| Bhatta | 60 | 04 01 20 | 07 30 20 | W India (Maharashtra) | 600 | 63.7 | m 42.9 | 56.2 | 13.3 |
| Panda | 61 | 4 23 20 | 06 29 20 | N India (Delhi) | 225 | 22.1 | m 35 | 70.7 | 0 |
| Chaurasia | 62 | 04 01 20 | 07 31 20 | N India (Uttarakhand) | 465 | 18.9 | N D | N D | N D |
| Malik | 63 | 04 29 20 | 07 20 20 | Bangladesh | 139 | 38.9 | m 34.1 | 41 | 1 |
| Shah | 64 | Apr 2020 | Aug 2020 | N India (Srinagar) | 655 | 18.5 | m 32.7 | 63.2 | 0 |
| Bhattacharjee | 65 | 05 12 20 | 05 21 20 | W India (Maharashtra) | 34 | 15 | m 35 | 61.8 | 0 |
| Jain | 66 | May 2020 | Jun 2020 | N India (Delhi) | 410 | 22.4 | m 37.2 | 63.9 | 0 |
| Dev | 67 | 05 01 20 | 06 15 20 | N India (Delhi) | 261 | 21 | m 36 | 58 | 0 |
| Kumar L | 68 | May 2020 | Aug 2020 | N India (Delhi) | 141 | 24.1 | m 15.2 | 58.9 | 0 |
| Koul | 69 | 05 15 20 | 08 15 20 | N India (Jammu) | 300 | 42.5 | m 37 | 74 | 0 |
| Sahoo | 70 | 06 25 20 | 07 24 20 | E India (Odisha) | 718 | 10.7 | m 34 | 78 | 0 |
| Rajkumar | 71 | 07 06 20 | 07 12 20 | S India (Tamil Nadu) | 230 | 26.9 | m 43.6 | 63 | 0 |
| Kumar V | 72 | Jun 2020 | Aug 2020 | N India (Delhi) | 200 | 34.0 | m 37.4 | 75.5 | 0 |
| Kandakure | 73 | Jun 2020 | Sep 2020 | W India (Maharashtra) | 200 | 8.5 | ~ 35 | 54.5 | 0 |
| Hasan | 74 | Jul 2020 | Sep 2020 | Bangladesh | 600 | 38.7 | m 50.2 | 64 | 0 |
| Hajare | 75 | Jul 2020 | Sep 2020 | S India (Karnataka) | 167 | 62.9 | m 39 | 54.5 | N D |
| Thakur | 6 | Sep 2020 | Oct 2020 | S India (Karnataka) | 250 | 71.6 | ~ 44 | 57.6 | 0 |
| Savtale | 76 | 10 01 20 | 10 15 20 | W India (Maharashtra) | 180 | 55.5 | m 37.8 | 33.4 | 0 |
| Gupta | 77 | Nov 2020 | Nov 2020 | N India (Rajasthan) | 98 | 86.5 | m 41.4 | 49 | 0 |
| Mangal | 78 | 11 16 20 | 11 26 20 | W India (Maharashtra) | 57 | 17.7 | ~ 35 | N D | 0 |
| Karthikeyan | 79 | Aug 2020 | Dec 2020 | S India (Tamil Nadu) | 1,000 | 74.2 | m 42.4 | 59.6 * | 0 |
| Silu | 80 | Sep 2020 | 01 15 21 | N India (Rajasthan) | 2,055 | 43.0 | m 50.4 | 77.4 | 2.8 |
| TOTAL |  |  |  |  | 9,626 | Mean= 31.8% | Mean= 34.32 | Mean= 61.35 | Mean= 1.64% |

Abbreviations: N D, not disclosed; \*, 3.2% transgender was assigned 50/50 to male and female gender.
Virus dominance was determined by multiple studies as described in the section "Spatiotemporal mapping of D614 or G614 virus dominance in South Asians"

The main novel finding of our meta-analysis is that, when the same ethnicity is compared for anosmia prevalence under two different conditions, with either the D614 or G614 virus strain being dominant, there is a large difference in anosmia prevalence between the two conditions. This means that the more infectious virus type, G614, is a significant factor for anosmia prevalence, apparently more important than ethnic differences in variants of the host proteins, ACE2 and TMPRSS2. This is presumably due to a higher efficiency of G614 virus entry to the host cell, ^11, 13, 81^ as discussed at the end of this review.

### Effects of response bias, age, disease severity, gender, and methodology

Are there alternative interpretations of our data that could explain, entirely or partially, the differences in anosmia prevalence among South Asian populations? Three types of parameters need to be considered: response bias, demographics, and methodology. If the demographics of the two cohort types (D614 vs G614 virus dominance) differed, they could have influenced or biased the comparisons. Demographics that have been shown or suspected in previous reviews to have an association with anosmia prevalence include age, gender, and COVID-19 disease severity. Age was associated in some analyses. ^1, 82^ Gender was found to show trends in the largest meta-analysis, ^1^ but not in two earlier analyses that considered fewer studies. ^82, 83^ Multiple studies and reviews agree that disease severity is negatively associated with anosmia prevalence. ^1, 83–85^ Finally, objective methodology to assess olfactory dysfunction may be more sensitive than subjective recall, resulting in an increased prevalence, although studies are controversial. ^7^ Response bias may occur if early studies did not inquire about olfactory dysfunction as much as later studies.

Response bias. Early in the pandemic, there was no publicity about COVID-19 causing loss of smell. Patients, caregivers and investigators may not have asked about this symptom or regarded it as irrelevant, resulting in response bias compared with later studies. This issue has been widely discussed. ^1, 2, 4, 5, 10, 83, 87, 88^ Could this bias explain why early studies in Asia did not report olfactory dysfunction as often as subsequent studies on Caucasians? This is unlikely to be a decisive factor for two reasons. Careful analyses of East Asians for chemosensory dysfunction revealed low prevalence, ^5, 89, 90^ even with objective olfactory testing, ^91^ and when the olfactory dysfunction was re-examined in Chinese populations at a later date, the anosmia prevalence still was significantly lower in Chinese patients than in Caucasian patients. ^92^ Second, we show here for South Asians that even studies conducted later in the pandemic, when the media had widely publicized and revealed anosmia/hyposmia as a cardinal symptom of COVID-19, reported a low prevalence when they examined cohorts in a region (e.g. Delhi and Karnataka) and at a time when the D614 virus strain was dominant. Therefore, we can exclude response bias as the main explanation for prevalence differences in South Asians.

Age. When we tested the parameter of age in our pooled analysis, we found no significant difference between the two groups (one group infected predominantly with the D614 virus, the other with the G614 virus). Mean age was 38.83 years +/- 1.23 standard error (SE) for the D614 group, and 34.32 years +/- 1.53 SE for the G614 group, with p= 0.3288 (β= 0.037 +/- 0.038 SE), which is no significant difference.

Disease severity. As a measure of COVID disease severity, we calculated the percentage of hospitalizations or percentage of cases with “severe” disease, when disclosed, and we found in our pooled analysis that the D614 group had a mean 5.85% severe cases +/- 3.82 SE, while the G614 group had 1.64% +/- 1.05 SE. The subgroup test showed no significant difference in anosmia based on disease severity with p= 0.5294 (β = −0.0173 +/- 0.027 SE). We conclude that disease severity does not explain the prevalence differences between the cohorts with D614 or G614 virus dominance.

Gender. It is unclear whether gender is associated with anosmia in COVID. ^1, 10, 82, 83^ We tested the parameter of gender using percent male in each cohort as a continuous variable in a meta-regression. In both groups, there were more males than females. Specifically, among the D614 group, there was a mean of 75.08 +/- 3.57% males, while there was a mean 61.35 +/- 2.45% males in the G614 group (mean +/- SE). The subgroup test was significant (p< 0.0001) with a negative linear relationship between percent of cohorts that were male and the prevalence of anosmia (β = −0.0586 +/- 0.0144 SE; Supplemental Fig. 1). Accordingly, a contributing effect of gender cannot be ruled out. However, the possible gender effect on anosmia prevalence is small: we calculated that the difference in the male/female ratio between the two cohort groups (Table 1) would account for less than 1/10th of the observed difference in anosmia prevalence between the two groups (5.3% vs. 31.8%, Fig. 3A).

Multivariable meta-regression. When we modeled age, gender, and group (D614 or G614 virus dominance cohorts) together, cohort remained highly significant at p<0.0001 (β= 1.95 +/- 0.363, D614= reference), while gender and age showed marginal significance at p=0.0549 and p=0.0484, respectively (β_gender_= −0.0235 +/- 0.0122, β_age_= 0.0477 +/- 0.0242). We did not include disease severity in our multivariable meta-regression due to missing data: 76.7% (n/N=33/43) of the studies provided data on all four variables whereas modeling only age, gender, and group allowed us to use 90.70% of the studies (n/N= 39/43) in our multivariable regression.

Methodology. Chemosensory dysfunction in COVID-19 can be assessed by questionnaires and history taking (subjective tests), or by testing the sense of smell objectively. ^1, 7, 93^ Some studies using objective tests showed an increase in the prevalence of olfactory dysfunction, ^1, 2, 82, 93–95^ while other studies reported the opposite (reviewed in Boscutti et al., 2021). ^7^ In the studies on South Asian COVID-19 patients, objective tests were used in only three of the cohorts with G614 virus dominance. To avoid methodology as a confounding variable, we considered only studies with subjective questioning. We can therefore rule out methodological parameters as an explanation for the difference in anosmia prevalence between D614 and G614 cohorts.

Taken together, we conclude that response bias, age, disease severity, and methodology cannot sufficiently explain the difference in prevalence between the two types of cohorts, and that after adjusting for the effects of age and cohort virus type, the effect of gender is relatively small compared to that of the cohort virus effect. Accordingly, given that we controlled for ethnicity by including only cohorts with South Asian populations, the virus type (G614 vs. D614) appears to be the most relevant parameter that is responsible for the vast majority of the observed prevalence differences.

### Limitations

Sampling of virus genomics differs between regions in South Asia, some regions have low sample numbers, and results may not be representative for all of these regions. Some studies reported only approximate dates for the beginning and end dates of study periods; furthermore, when data on anosmia were collected over a longer period of months, it is possible that the majority of patients were enrolled early or late during that period, making the correlation with the virus strain less precise. We cannot entirely rule out a small contribution of response bias – earlier studies may have insisted less on information about olfactory dysfunction than later studies. Finally, there is some heterogeneity, possibly due to yet unknown parameters and lack of information about disease severity among patients within cohorts.

### Supporting evidence for a role of G614 in anosmia prevalence

Our meta-analysis of studies on South Asians provides a strong case for a role of the D614G virus mutation in anosmia prevalence. Is there additional evidence from studies on other ethnicities to support this notion? The study by Eyre et al. (2020) ^96^ examined anosmia prevalence in the UK separately for Caucasians, Asians, and Chinese subjects residing in the UK, and found no significant difference in prevalence between ethnicities at a time when the G614 virus was predominant. This supports the idea that ethnicity plays no or only a minor role in anosmia prevalence, although the data on “Chinese” was based on a very small cohort (n=10).

The prevalence of anosmia was also examined by three studies in Hong Kong, ^97–99^ at a time when the G614 virus began to dominate over the D614 virus (March-April 2020). ^14,100^ Apparently, the G614 virus was dominant in Hong Kong because of the extensive travel to Hong Kong from the UK and USA ^99^ where the G614 virus had taken over. This region is the one exception to the predominance of the D614 virus throughout the rest of mainland China in 2020. ^14^ All three of the studies in Hong Kong found the prevalence of anosmia to be much higher (66.7%, 47.0%, and 22.1%, – weighted mean = 37.44%) than from other regions in East Asia (pooled mean of 16.7%), ^1^ although one of the studies from Hong Kong reported on a very small cohort (n=18). ^97^ The studies from Hong Kong again point to the G614 virus as the reason for the increased anosmia prevalence. When anosmia prevalence was determined early during the pandemic in Malaysia (in April 2020), while the D614 virus was still dominant, with only a minor contribution of the G614 virus, the anosmia prevalence was 21.4%. ^37^ Just 3-5 months later, when the G614 virus dominated, ^14, 38^ the anosmia prevalence had increased to 36.6%, ^101^ consistent with our hypothesis.

There are some other indications, from regions in Europe, that a larger fraction of the D614 virus early in the pandemic may have contributed to a relatively low anosmia prevalence, e.g., in Iceland ^102^ and in Spain. ^103^ However, while suggestive, the period of data collection in these studies was at a time when the two virus strains co-existed, making it impossible to know whether the patients with low anosmia prevalence were indeed mostly those infected with the D614 virus. Furthermore, the study from Spain ^103^ primarily examined hospitalized patients, and disease severity associates negatively with anosmia prevalence, as mentioned above. Taken together, the Hong Kong studies ^97–99^ provide the most convincing supporting evidence outside of South Asia for a contribution of the G614 virus to anosmia.

### Is there also a contribution of human host variants – true ethnic differences?

There are several variant proteins in the human host which could cause ethnic differences in anosmia prevalence. The one that has been most discussed is ACE2, which has variants that are known to differ in their binding affinities to the virus, or their methylation status, ^104^ and the frequency of such variants differs between ethnicities. ^104–, 108^ Another protein is the protease TMPRSS2 which cleaves the spike protein, allowing fusion and cell entry, and there are variants of TMPRSS2 that differ between ethnicities. ^109, 110^ Virus properties affected by the D614G mutation may further have downstream effects on virus entry because of ethnically distinct differences (e.g., in the frequency of the alpha anti-trypsin protease inhibitor). ^111^ Recently, a third category of genes has been found to differ between ethnicities. ^112^ The odorant metabolizing enzyme UGT2A1 which is expressed primarily in sustentacular cells and olfactory cilia ^113^ was associated with COVID-related anosmia in a trans-ethnic analysis. ^112^ Such studies support the idea that host protein variants may contribute to the extent of anosmia and could explain, at least in part, ethnic differences in anosmia prevalence in COVID-19 (reviewed in Butowt et al., 2020). ^8^

When using the same meta-analytic methods to compare cohorts primarily infected with the same virus strain (thus controlling for the virus type at a coarse level), the prevalence of olfactory dysfunction in the South Asian population was significantly lower than the prevalence reported for Caucasians (p= 0.0054, Fig. 4B, D). The weighted random prevalence of anosmia among Caucasian COVID patients was 49.02% (95% CI= 42.25-55.84%, N=77) and 31.79% (95%CI= 23.26-41.76%, N=25) among South Asian COVID patients. This suggests a relatively small, but measurable contribution of the host (ethnic difference in frequency of ACE2 or proteins such as UGT2A1).

### Why does SARS-CoV-2 cause much more anosmia than SARS-CoV-1?

The mechanism by which SARS-CoV-2 causes anosmia is beginning to come into view. Several different scenarios were initially considered: nasal obstruction (possibly due to inflammation), olfactory receptor neuron damage, olfactory support cell damage, and damage to central olfactory pathways. ^3, 4, 10, 114, 115^ An emerging consensus favors a crucial role of the sustentacular support cells in the olfactory epithelium as the primary mechanism of COVID-induced anosmia. ^5, 8, 114, 116–120^ Since the SARS-CoV-2 entry protein (ACE2) is not or only minimally expressed in olfactory receptor neurons, the virus rarely infects the neurons, ^121^ but rather enters the olfactory epithelium through sustentacular support cells and secretory cells in Bowman glands and primarily damages these cell types (Fig. 5A). When sustentacular cells become infected, they rapidly die, which appears to cause retraction of the adjacent neurons’ ciliary processes, ^116, 122^ and may down-regulate expression of odorant receptors necessary for olfactory transduction. ^123^ In addition, infection and damage of Bowman gland cells may alter the composition of the mucus that is required for efficient access of odorants to the odorant receptors on the neuron’s cilia. ^119, 120^ This prevents or alters binding of odorants to the olfactory neurons and thereby impacts olfactory transduction. After the degeneration of the sustentacular cells, stem cells in the olfactory epithelium divide (within a few days after death of sustentacular cells) and rapidly regenerate the lost sustentacular support cells, ^114^ allowing the olfactory epithelium to be repaired. The neurons then recover, restore their cilia, resume odorant receptor expression, and the sense of smell returns in most cases within 1-2 weeks. ^1^ One study has proposed that the SARS-CoV-2 virus resembles olfactory receptors, and that IgA produced against the virus may thereby compromise olfaction as “collateral damage.” ^124^ However, the early timing of loss of smell vs. the delayed production of IgA makes this model an unlikely mechanism, although antigen exposure via the olfactory epithelium may indeed boost the immune response and could lead to a more successful viral clearance and milder COVID-19 at the cost of a (temporary) loss of smell. ^120^

**Fig. 5A. B.**
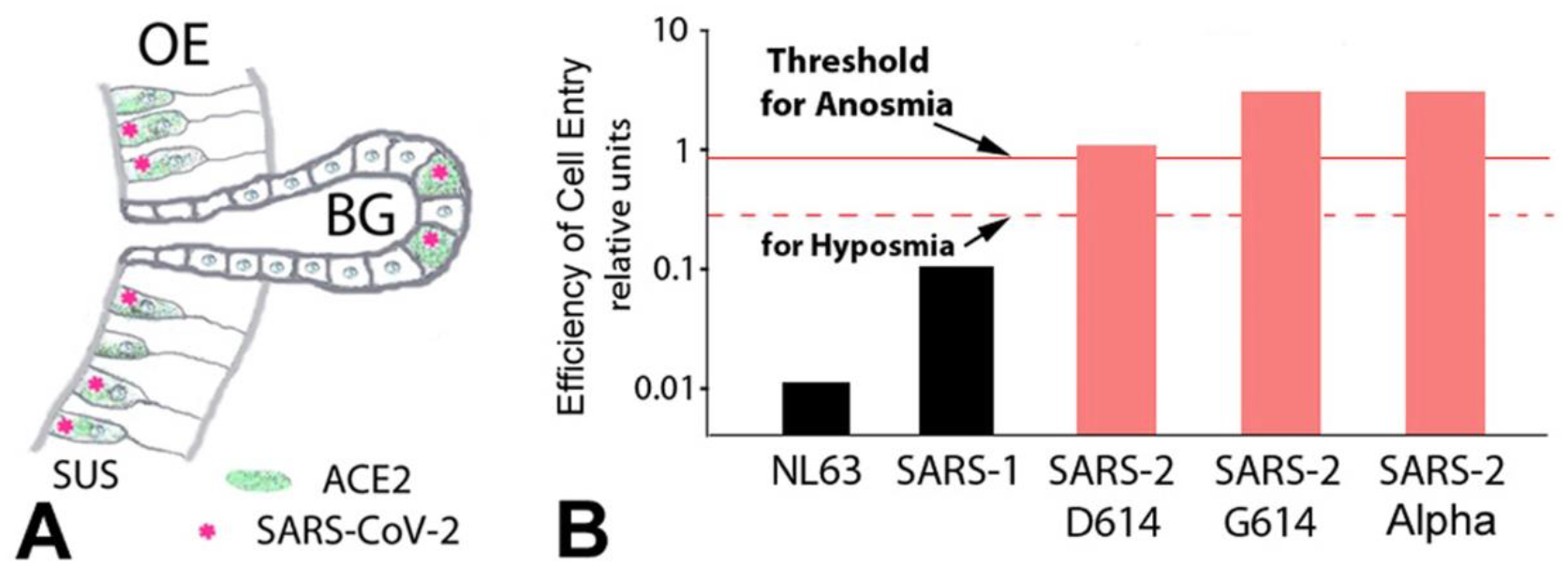
Illustration of the cell types infected in the olfactory epithelium (OE) (panel A) and the concept that the cell entry efficiency of the coronavirus determines the extent of damage that causes hyposmia or anosmia (panel B). This would explain the varying prevalence of olfactory dysfunction in patients with COVID-19 between populations and dominance of human coronaviruses NL63, SARS-1, SARS-2, and its strains D614, G614 or the G614 Alpha variant. Efficiency of cell entry includes differences in binding affinities, fusion efficiency via the novel furin cleavage site, and neuropilin-1 binding as a co-host. For details, see Butowt et al., 2020. ^8^ ACE2, angiotensin converting enzyme 2; BG, Bowman gland; SUS, sustentacular cell.

While the SARS-CoV-2 virus readily induces anosmia, the SARS-CoV-1 virus does not, even though the two virus types bind to the same entry receptor, ACE2. ^8^ So what is different? The SARS-CoV-2 virus has a significantly higher binding affinity to the ACE2 receptor than the SARS-CoV-1 virus (Fig. 5B). In addition, the SARS-CoV-2 virus has a furin cleavage site that SARS-CoV-1 does not have. ^125^ By enhancing fusion efficiency, this site is thought to make SARS-CoV-2 more pathogenic. ^126^ The furin cleavage site may increase tropism (widening of the range of susceptible host cells) and cause higher pathogenicity. ^109, 127^ The higher binding affinity and the new furin cleavage site may both be responsible for the higher infectivity and higher anosmia prevalence of SARS-CoV-2 compared with SARS-CoV-1. Cleavage of the S1 spike protein generates a neuropilin-1 binding site, and therefore neuropilin-1 can act as a host factor for SARS-CoV-2. ^128, 129^ Neuropilin is enriched in the olfactory epithelium, ^128^ indicating that the increased anosmia may involve binding to neuropilin-1.

### The role of the D614G mutation for anosmia – presumed molecular mechanisms

A key finding to understand how SARS-CoV-2 causes anosmia is that the mutated G614 virus is much more detrimental to olfaction than the original D614 virus. Why is the D614G mutation so much more effective in targeting the olfactory epithelium? What are the molecular mechanisms of this mutation? A number of studies have explored consequences of the D614G mutation, both for the clinical phenotype and for the pathophysiology at the molecular level. Clinically, despite the higher viral load, studies failed to detect an effect of the D614G mutation on the severity of COVID-19, hospitalization rate, or mortality. ^14, 130, 131^

Regarding the molecular mechanism of how the D614G mutation increases infectivity, transmission and possible disease severity, four different mechanisms are currently discussed, as recently reviewed. ^132–134^ These are (1) modulation of the spike protein (by adding an elastase cleavage site and/or making furin cleavage more efficient); ^135, 136^ (2) promoting an open conformation of the receptor binding domain that favors ACE2 interaction; ^13, 81, 137^ (3) increasing spike density and therefore facilitating cell entry; ^81^ and (4) enhancing the stability of the spike protein (stronger retention of S1, less shedding of S1). ^138^ All four mechanisms may contribute to enhanced cell entry and infectivity of the G614 virus.

But does this explain why the G614 virus is so much more effective at attacking the olfactory system? The G614 virus infects the upper respiratory tract, including nasal epithelium, more than lung epithelial cells, resulting in higher viral loads in the olfactory epithelium than in the lower respiratory tract. ^117, 139^ The enhanced cleavage at the furin cleavage site is of potential interest, because the support cells and Bowman gland cells in the olfactory epithelium co-express not only ACE2 and TMPRSS2, but also furin. ^140^ Accordingly, the cells in the olfactory epithelium may be more efficiently infected with the G614 virus, and this may explain the higher viral load in the nasal epithelium than in the lungs. ^111^ If the replicating virions are already cleaved by intracellular furin, as they exit the host cell, then they are ready to fuse with the next host cell, resulting in a more fulminant spread. ^141^

On the other hand, the G614 mutation exposes the virus spike protein more than the D614 virus does, making the G614 virus more immunogenic, and possibly eliciting a stronger host immune response. ^11, 120, 137, 141^ Such differences in immunogenicity may indicate that the G614 virus, when it infects the nasal epithelium with a higher viral load, may trigger a more robust host immune defense. Although the virus may move to a new host too quickly to be affected by a neutralizing antibody response, ^134^ it cannot be ruled out a scenario where the virus in the olfactory epithelium (often preceding any other symptoms) elicits an immune host response that gives the host enough time to accelerate virus clearance and to prevent a subsequent more deadly infection of the lungs. ^120^ This may explain why SARS-CoV-2 infection leading to anosmia is associated with an overall milder COVID-19 disease, possibly because the nasal cavity-elicited immune defense leads to faster virus clearance ^143^ and thus reduces severe COVID-19 lung disease and death after G614 virus infection. ^144, 145^ Such a scenario may explain the puzzling finding that the G614 virus, despite being more infectious and leading to higher nasopharyngeal viral loads than the D614 virus, does not – overall – cause more severe and deadly COVID-19.

The presumed mechanisms of increasing efficiencies of binding and cell entry are summarized in Fig. 5A, B. Regardless of the precise mechanism, our analysis shows an increased prevalence of olfactory dysfunction in cohorts infected with the G614 virus. Apparently, the G614 virus is more efficient than the D614 virus in entering and damaging the olfactory epithelium and impairing olfactory function. Importantly, our review identifies a “missing link” by revealing increased prevalence of chemosensory dysfunction as a novel, previously unrecognized phenotype of the now dominant G614 virus.

### How will current and future novel virus variants affect olfactory function?

Since the spread of the G614 virus throughout most of the world, several new SARS-CoV-2 virus variants have emerged – Alpha (B.1.1.7), Beta (B.1.351), Gamma (B1.1.28), and Delta (B.1.617.2). All of these new variants also harbor the D614G mutation, ^4^ and therefore can be expected to cause similarly increased olfactory dysfunction as the G614 virus. These variants have additional spike protein mutations, besides the G614 mutation, resulting in already proven or suspected differences in their receptor binding properties, transmissibility, viral loads, and, in some cases, increased mortality. ^146^ For example, the virus with the N501Y mutation has similar binding to ACE2, while the K417N and the E484K mutants may have slightly increased binding to ACE2. ^147^ Multiple mutations can have interdependent and complex effects on binding and subsequent steps such as membrane fusion and host cell entry. For example, the Alpha variant that often also has N501Y, N439K and Y453F mutations appears to require a deletion (Δ H69/V70) in the spike protein to maintain optimal cleavage and infectivity. ^148^ How these mutations generally affect infectivity in vivo, and specifically for cells in the olfactory epithelium, is not yet known.

The prevalence of olfactory dysfunction has been reported so far in only one of these variants (Alpha); it did not cause a significant change in anosmia prevalence. ^149^ An additional commentary that was based on apparently less reliable data reported an anosmia prevalence for the Alpha variant that differed by less than 4% from that of the G614 virus. ^150, 151^ Future studies will be needed for reliable and conclusive data on the phenotypes of these new variants in terms of ACE2 binding affinity, membrane fusion, spike protein shedding, efficiency of host cell entry, viral load in different tissues, transmissibility, infectivity, mortality, and chemosensory dysfunction. Such phenotypes cannot be reliably predicted based on atomic modeling of the receptor binding domain of the spike protein, due to the assumptions and restrictions of the modeling parameters. ^152^ The cell entry properties of the new virus variants likely will continue to cause chemosensory dysfunction, and otorhinolaryngologists should expect to see such symptoms in COVID-19 patients when the new variants take over, as long as they maintain the D614G mutation, and assuming that the additional mutations do not neutralize their apparent effect on olfaction.

## Data Availability

All data referred to in the manuscript are publicly available.

## Supplemental Figures

Supplemental Fig. 1: Bubble plot showing details of the subgroup comparison with a gender effect on anosmia prevalence (Suppl Fig. 1 - Bubble plot of correlation of gender and anosmia.pdf).

Supplemental Fig. 2: Funnel plot for prevalence of anosmia in D614 and G614 cohorts of South Asians with COVID (Suppl Fig. 2 - Funnel Plot South Asian Studies.pdf).

Supplemental Fig. 3: Funnel plot for prevalence of anosmia in G614 cohorts of Caucasians with COVID (Suppl Fig. 3 - Funnel Plot of Caucasian and South Asian Studies.pdf).

## Acknowledgments

The authors thank the following colleagues for insightful discussions: Bing Chen and Sandeep Datta (Harvard, USA), Thomas Hummel (University of Dresden, Germany); Bette Korber (Los Alamos National Laboratory), Gannon Mak (Dept. of Health, Hong Kong, China), Dennis Mathew (University of Nevada, Reno), Nicolas Meunier (University Paris-Saclae, France), and Mauricio Ponga (University of British Columbia, Canada). We also thank the following colleagues for providing additional information: Nitesh Gupta (Safdarjung Hospital, Delhi), Ankit Khurana (Hospital Rohini, Delhi), Vinod Kumar (All India Institute of Medical Science, Delhi), Nikitha Pillai (Kims Hospital, Bangalore), and Vishav Yadav (Patiala, Punjab).

Grant support: GM103554 from the National Institutes of Health (C.S.v.B.), and the “Excellence Initiative-Research University” programme at the Nicolaus Copernicus University (R.B.).

## Methods

For mapping and quantification of the contributions of D614 and G614 viruses in countries, we relied on the tracking website (https://cov.lanl.gov/apps/covid-19/map/) ^14^ and for the regional geographical resolution within and outside of India we consulted multiple sources ^14, 16–22, 24–29, 31–36, 100, 153^ Our study followed the PRISMA guidelines for systematic reviews and meta-analyses. ^41^ We searched the COVID-19 portfolio of the National Institutes of Health (https://icite.od.nih.gov/covid19/search/) with the key words „anosmia” or „smell” and “India” or “Bangladesh” or “Pakistan” on and before July 22, 2021, resulting in 598 records. In addition, PubMed was searched (“anosmia”, “COVID”, “India,” or “Bangladesh” or “Pakistan”), resulting in 104 records. After removal of duplicates, 351 full-length texts were screened for the inclusion criteria: South Asian ethnicity (Indian, Pakistani or Bangladeshi); a confirmed COVID-19 diagnosis; majority of subjects adults or teenagers; information about the weeks or months of the study period(s); subjective (not objective) testing for olfactory dysfunction; we accepted no case reports, and included only reports of primary data, but no reviews. Studies were excluded because of lack of subjective testing, ^154^ consideration of only severe COVID cases, ^155^ mostly East Asian rather than South Asian ethnicity, ^156^ or lack of necessary information about study periods despite repeated requests. ^157^ The studies that met our inclusion criteria typically were cross-sectional, retrospective, observational studies that could be affected by recollection bias. However, such bias would be expected to be of a similar magnitude in studies examining patients infected with the D614 virus and studies examining patients infected with the G614 virus. We found 40 studies reporting on 43 cohorts that met our inclusion criteria. We compiled olfactory dysfunction regardless whether taste was also affected or not. The included studies are listed chronologically and sorted by D614 or G614 dominance in Table 1. A pooled analysis was performed for prevalence, and significance and confidence intervals were calculated in the software R, version 3.6.1 (R Foundation for Statistical Computing, Vienna, Austria). To calculate estimates of pooled prevalence and 95% confidence intervals, we used the R-meta package, version 4.9-5, and the metaprop function. We used random effects models with the inverse variance method for pooling and the logit transformation for proportions. ^158^ For ease of interpretation, we back transformed and rescaled proportions to events per 100 observations. Subgroup analyses were conducted for binary group variables (D614 vs G614 and Caucasian vs South Asian) using the byvar statement of the metaprop function. All other subgroup tests used continuous variables and the metareg function, including the multivariable meta-regression. The subgroup age was a created variable that used the center of the sample, either the mean or the median, to mark the center of the age distribution, with the majority of studies reporting mean age (75.4%, N=32/43). Analysis of the heterogeneity across studies was done using the Maximum-likelihood estimator, Higgins’ I^2^ and Cochran’s Q method. ^158, 159^ Publication bias was assessed by visual inspection of funnel plots ^160^ (Supplemental Figs. 2 and 3). In all cases, significance was defined at α = 0.05.

## Author contributions

All authors contributed to the conception and design of the manuscript.

## Supplemental Figures

**Supplemental Fig. 1:**
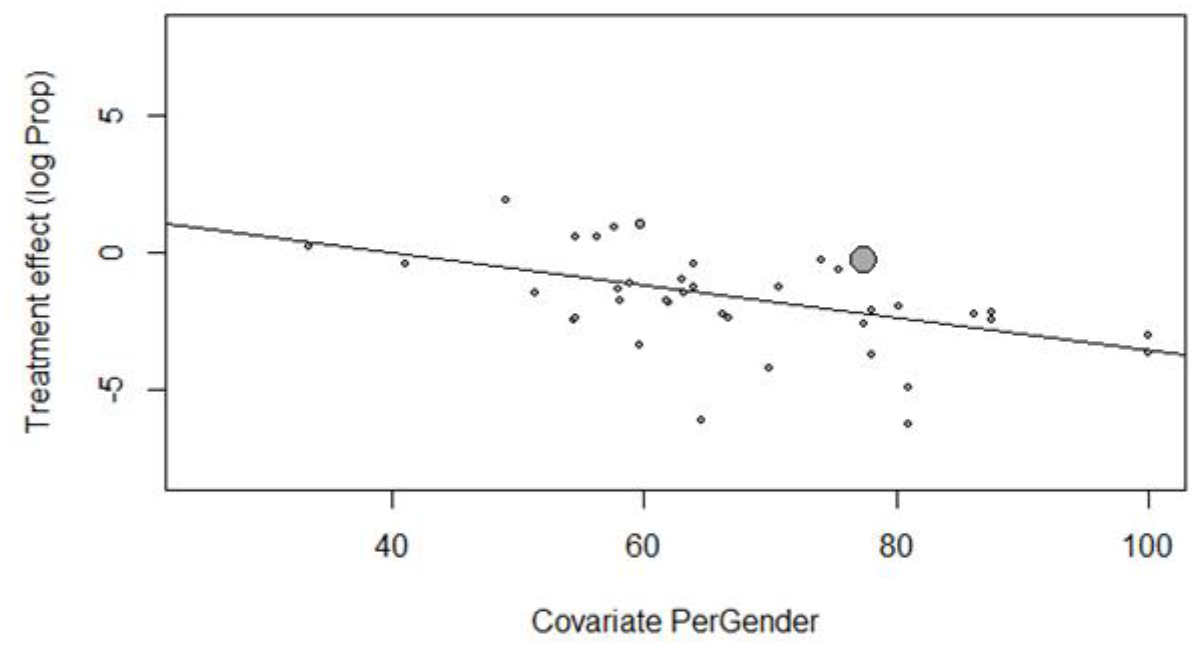
Bubble Plot showing negative correlation of male gender and prevalence of anosmia in south Asians.

**Supplemental Fig. 2:**
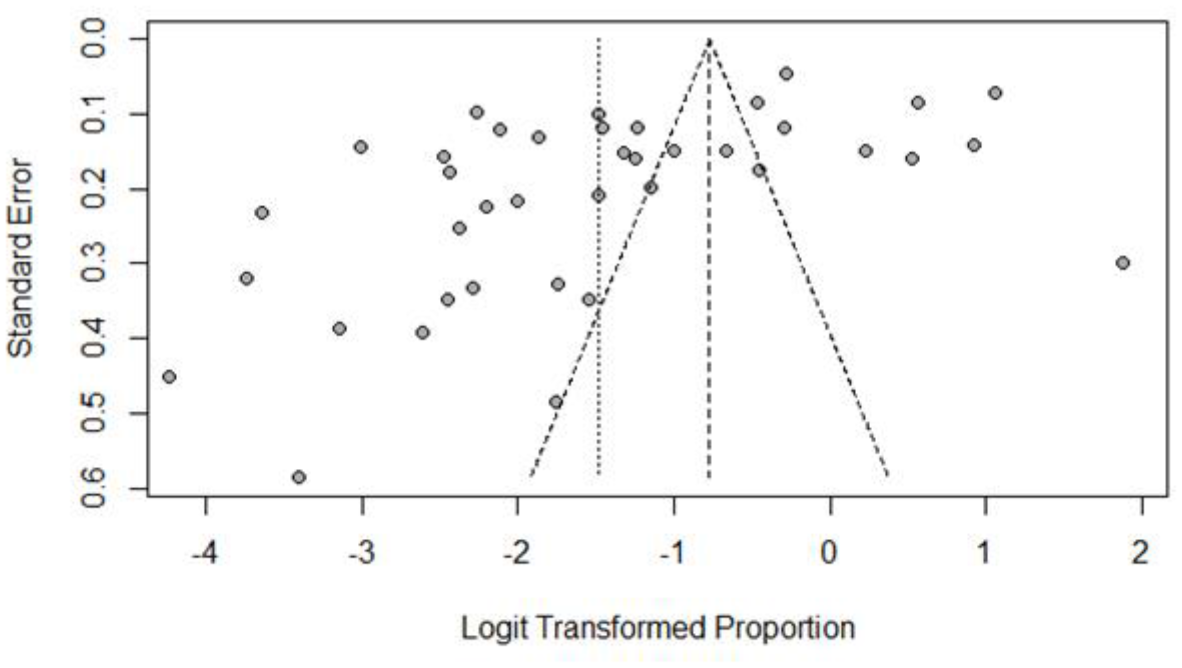
Funnel Plot of South Asian Studies (3 outliers removed)

**Supplemental Fig. 3:**
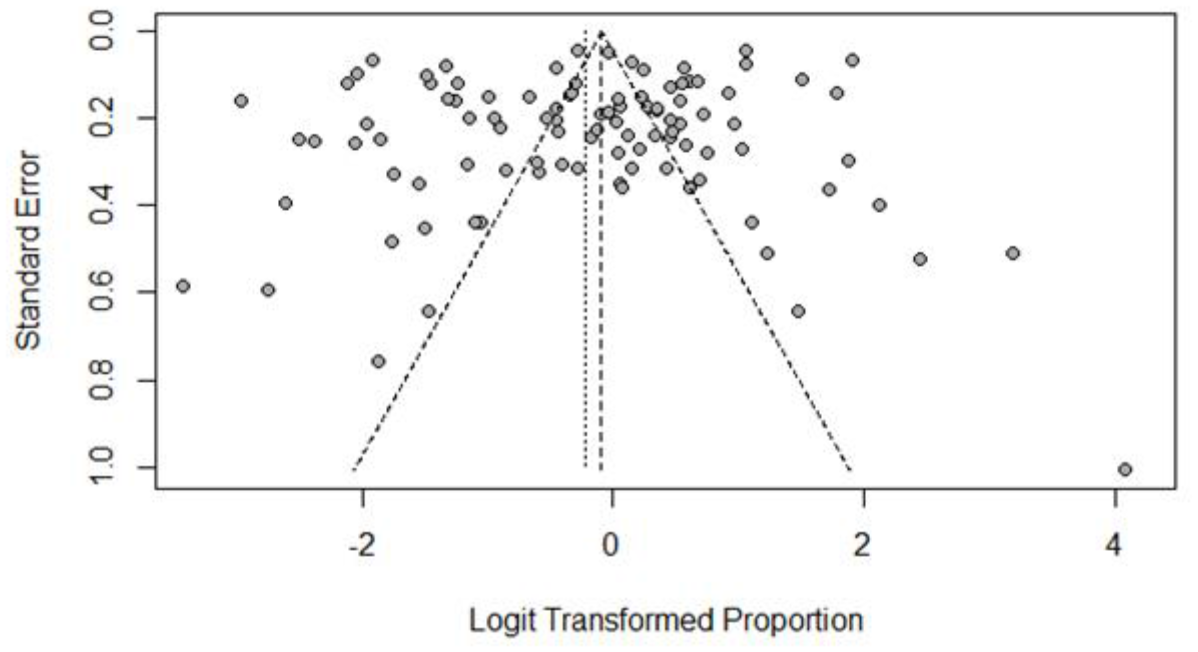
Funnel Plot of Caucasian and South Asian Studies

